# Potassium Reference Values in Neonates: Impact of Sampling Method and Clinical Condition

**DOI:** 10.64898/2025.12.12.25342135

**Authors:** Rudolf G. Ascherl, Benjamin W. Ackermann, Matthias Knüpfer

**Affiliations:** Department of Neonatology, Hospital for Children and Adolescents, Leipzig University Medical Center, Leipzig, Germany

## Abstract

**Background and Objectives:** Current potassium reference intervals for neonates fail to account for sampling method differences and prematurity-related factors, leading to unnecessary resampling and interventions. We aimed to establish sampling-specific potassium reference intervals for term and preterm neonates using comprehensive electronic health record data.

**Methods:** We analyzed 195 606 blood gas measurements from 10 290 neonates (2007-2024) at a tertiary neonatal intensive care unit. After ex-cluding values during severely impaired clinical conditions using integrated clinical metadata, we derived reference intervals for venous, arterial, and cap-illary samples. Multivariate analysis identified factors affecting potassium homeostasis.

**Results:** From 55 664 included values, capillary samples showed signif-icantly higher potassium levels than arterial or venous samples. Reference intervals (2.5^th^-97.5^th^ percentiles) for neonates >7 days: venous [2.6, 5.5] mM, arterial [2.6, 5.9] mM, capillary [3.0, 6.3] mM. Time-matched analysis of 5403 paired samples showed capillary-specific intervals achieved 88% sensitivity and 94% specificity for detecting true hyperkalemia compared to arterial or venous controls.

**Conclusion:** Capillary blood gas potassium levels require distinct, higher reference intervals than venous or arterial samples in neonates. Implementation of sampling-specific reference ranges may reduce false-positive results and unnecessary interventions in this vulnerable population.

**Article Summary:** Large EHR-based study defines sampling-specific neonatal potassium reference intervals; higher capillary ranges reduce false positives and maybe unnecessary interventions.

**What’s Known on This Subject:** Existing neonatal potassium reference intervals often ignore sampling modality and prematurity, contributing to clinical uncertainty and unnecessary repeat testing in neonates.

**What This Study Adds:** Provides sampling-specific potassium reference intervals for neonates; capillary samples require distinct, higher ranges, improving discrimination of true hyperkalemia and reducing false positives.

**Contributors Statement Page:** **Rudolf G. Ascherl:** Conceptualized and designed the study, coordinated data extraction, perfomed analysis and visualization, drafted and revised the initial manuscript.

**Benjamin W. Ackermann:** Contributed to study design, assisted in revising the manuscript.

**Matthias Knüpfer:** Provided clinical oversight, interpreted findings, critically reviewed the manuscript.

*Equal contribution:* Rudolf G. Ascherl and Benjamin W. Ackermann.

**All authors approved the final manuscript as submitted and agree to be accountable for all aspects of the work.**

## Introduction

Reference values are indispensable for interpreting laboratory results. They are given as a cut-off or an interval estimate spanning 95 % of a reference population. [1]

While clinicians caring for adults can easily rely on blood donors, military re-cruits, or healthy subjects of clinical studies as reference populations, the pediatric population has no such groups. That is because in children there are ethical and practical barriers to the establishment of such cohorts.[2] Usually in the adult, the only distinction in reference values is made regarding sex; in the pediatrics mat-urational changes such as growth, puberty and, specific to the neonate, postnatal adaption need to be addressed. This would make much larger reference samples necessary complicating the aforementioned problems of recruiting a reference population.

Notwithstanding very few examples mentioned later on, studies into the topic of RI have at best pushed these boundaries: The studies KiGGS and LIFE Child did include neonates, however, no blood was sampled in in KiGGS during the first year of life and in LIFE Child not before 3 months of age. [3, 4]

Preterm birth entails further complications: A sample volume of for example 200 µL is not just practically difficult to obtain in a premature infant weighing 0.5 kg – if repeated more than 3 times it is also in excess of 1 % of the patient’s total blood volume. To add to that, fewer than 1 % of all newborns have a birth weight below 1.5 kg, so there are few patients to study especially in relation to the larger sample size needed to represent maturation. Challenges in drawing blood frequently compel neonatologists to resort to capillary sampling. That short gestation and low birth weight are recognized as diagnoses and that they indicate intensified care adds to that; preterm infants are thus not *healthy* by common definitions. Cord blood reference values at birth have been brought forward, but changes due to adaption to extrauterine live limit their postnatal use. All of this ends in a lack of reference values in neonatology, which has been recognized as a detriment at the very least to research purposes. [5]

With the recognition of its long-lasting effects on the patients, pain reduction has become a priority in the care of preterm infants. The high temporal dy-namic and uncertainties regarding the normal clinical course, make comparisons of repeated laboratory results necessary. This tempts physicians to request labo-ratory investigations in this very vulnerable and relatively new patient group more frequently.

Just like a blood donor will not be sampled for reference values when he is sick, we aimed at excluding the potassium values sampled during severely reduced condition by means of a surrogate created from other values in our database of electronic patient records. We did this to arrive at a sample of laboratory results taken from relatively healthy preterm neonates. From this sample we want to derive reference intervals (RI) pertinent to the term and preterm infant population and gain further insight into the postnatal course of potassium. Such specific reference values may aid in reducing blood sampling and the pain and anemia that arise from it in this extremely vulnerable group.

## Methods

### Patients and Setting

Data for this study were extracted from routine electronic patient records in our tertiary neonatological intensive care unit and include values acquired between 2007 and 2024. The patient data management system automatically recorded a mean value of vital signs and ventilator data every 15 minutes. Nurses entered weights, medications, and outputs manually into the same patient data management system; output was either derived by weighing diapers or, if a urinary catheter was present, read from the collector. We selected only those admitted until their 2^nd^ day of life to rule out treatment differences in those outborn. Potassium was measured via ion-selective electrodes in ABL 800 FLEX blood gas analyzers (Radiometer, Brønshøj, Denmark). [6] The institutional review board at the Medical Faculty of the University of Leipzig reviewed the protocol and waived consent (internal reference number 270/23-ek).

### Filters, Annotation and Sampling Strategy

A database of all laboratory results was filtered for potassium values from blood gas analyzers after stripping duplicates. These values were then annotated with the most recent weight of the patient, sex, gestational age, and postmenstrual age at sampling.

We filtered out all values at which potassium is life threatening by itself, i. e. *K* < 1.5 mM or *K* > 10 mM.

To obtain a subset of samples taken predominantly from healthy patients, we aimed at excluding all values sampled during phases of severely impaired condition (SIC). In brief, to be of SIC, at least one of the following had to be present each within their own specific time frame before sampling: high fraction of inspired oxygen (FiO_2_), low urine output, an increased difference of peripheral and central temperatures, or application of certain medications (e. g. inotropic or vasoconstrictive agents, doxapram, and buffers). Details of the SIC definition are described in table 1.

**Table 1:** Definition of the surrogate for severely impaired condition. Each of the four criteria was analyzed over a different time frame given as hours before sampling (h.b.s.). When at least one of the conditions was fulfilled the sample was considered as being taken during a severely impaired condition. * during 1^st^ day of life, this was corrected for time since birth

| Criterion | Time |
| --- | --- |
| **Oxygen** maximum fraction of inspired oxygen $\max(\text{FiO}_2) > 0.4$ | 1 h.b.s. |
| **Urine output** average urine output $\overline{UO} < 0.5 \text{ ml kg}^{-1} \text{ h}^{-1}$ | 24 h.b.s.* |
| **Temperature gradient** maximum difference between core and peripheral temperature $\max(\Delta T) > 2^\circ\text{C}$ | 4 h.b.s. |
| **Medication** any of the following medications: epinephrine, norepinephrine, dobutamine, adenosine, hemopressin, milrinone, sildenafil, sodium bicarbonate (excluding oral route), tromethamine, pentoxifylline, sodium benzoate, rasburicase, urokinase, alteplase, doxapram, surfactant, recombinant factor VIIa | 12 h.b.s. |

In addition we flagged all samples for the use of the diuretics Furosemide and Spironolactone, 12 hours before sampling.

All of this allows to account for many aspects pertinent to potassium homeostasis, such as respiratory acidosis, renal function, and medication.

### Statistics

All values are given as Median [2.5^th^, 97.5^th^ centiles] unless denoted otherwise. Analyses were conducted in the R software environment. Densities were estimated using Gaussian kernels. Preliminary analysis using the direct plug-in method by Sheather and Jones [7] and Silverman’s rule of thumb [8] yielded bandwidths within ±15% of the precision of the blood gas analyzer of 0.1, which we thus as-sumed for all kernel density estimations. Since all distributions in this preliminary analysis were monomodal, we assumed the mode *x̂* to be at the highest density. We used the `HDInterval` package to calculate the credible intervals *CrI* with a mass of 95 %. [9]

## Results

### Population Characteristics

Of all patients treated at our NICU at the aforementioned time interval 10290 neonates had at least one blood gas analysis and were thus included in this study. Their characteristics are summarized in table 2.

**Table 2:** Population characteristics. Fractions for BPD, NEC, SIP, IVH, and ROP are in relation to children with a gestational age below 32 weeks; others are related to *N*. BGA, blood gas analysis; BPD, bornchopulmonary dysplasia; IVH, intraventricular hemorrhage; NEC, necrotizing entercolitis; ROP, retinopathy of prematurity; SDS, standard deviation score; SIP, spontaneous intestinal perforation

| Variable | Value | N/A |
| --- | --- | --- |
| N | 10290 | - |
| birth weight [kg] | 2.56[0.69, 4.29] | 153 |
| birth weight [SDS] | -0.13[-2.37, 1.94] | 155 |
| gestational age [d] | 255[179, 290] | 0 |
| moderate preterm | 3692 (35.9%) |  |
| very preterm | 1302 (12.7%) |  |
| extremely preterm | 660 (6.4%) |  |
| APGAR 5' | 8[5, 10] | 282 |
| APGAR 10' | 9[6, 10] | 291 |
| umbilical artery pH | 7.29[6.98, 7.42] | 828 |
| first hospital stay [d] | 10.9[0.4, 97.2] | 19 |
| BPD<sub>O<sub>2</sub></sub> at 28 d | 655 (12.2%) | - |
| BPD<sub>36 weeks</sub> | 153 (8.19%) | - |
| NEC | 33 (1.77%) | 290 |
| SIP | 77 (4.12%) | 234 |
| IVH > 1° | 210 (3.9%) | 658 |
| ROP > 1° | 212 (11.34%) | 8506 |
| treatment for ROP | 54 (2.89%) | - |
| death | 181 (1.76%) | - |
| BGA per patient | 8[1, 108] | - |
| BGA per patient and day | 0.77[0.18, 9.74] | - |

### Sampling Results

Numbers of samples at each step of the filtering, annotation, and exclusion are depicted in figure 1. The size of this indirect reference sample is 56 950, which is 29.8 % of the unique BGAs. Most values were filtered out due to SIC. More specifically, 51 % patients with arterial samples had all values filtered out by this; this fraction is 49 % for venous and 27 % capillary respectively.

**Figure 1:**
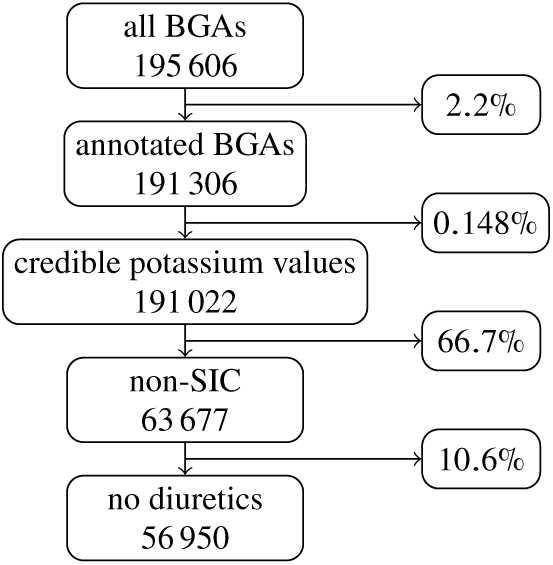
Flowchart of the laboratory value filtering process. The number below each stage denotes the remaining sample size, the percentages on the right are the fraction of the values excluded at each step. BGA, blood gad analysis; SIC, severely impaired condition those can be found in table 3. Apart from potassium intake, meaning additional potassium given in addition to parenteral or enteral nutrition, all variables had significant influence on potassium concentrations. Capillary sampling exhibited the most meaningful impact. Chronological and gestational ages showed much weaker, but yet significant, effects by comparison. The calendar date at which the sample was drawn was incorporated to model secular trends, reflecting systematic changes in practice patterns and external influences over the study period.

**Table 3:**
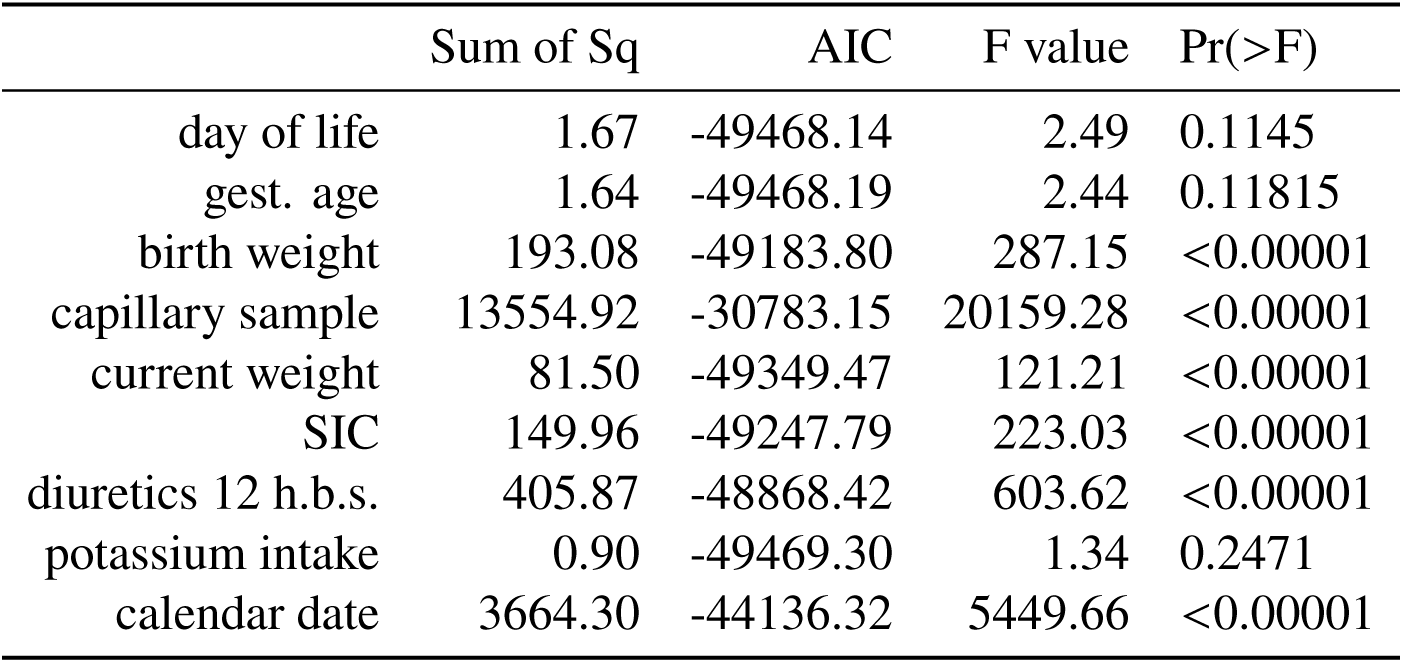
Single-term deletion model of potassium values. h.b.s., hours before sampling; gest. age, gestational age; SIC, severely impaired condition

The resulting sample is composed of 6% arterial, 17% venous, and 77% capillary BGAs.

### Multivariate Analysis

To test the validity of our exclusion strategy and identify further important vari-ables we conducted regression models with single term deletions. The results of

### Influence of Sampling Modality

Figure 2 displays the kernel density estimation of the distributions from each sampling modality and table 4 presents descriptive statistics: All three distributions exhibit positive skewness. The locations of the arterial and venous distributions are nearly identical, while the capillary exceeds them markedly. The scale is smallest for the arterial distribution, followed by the venous with the capillary having the highest scale; these differences, however, become very small if corrected for location.

**Figure 2:**
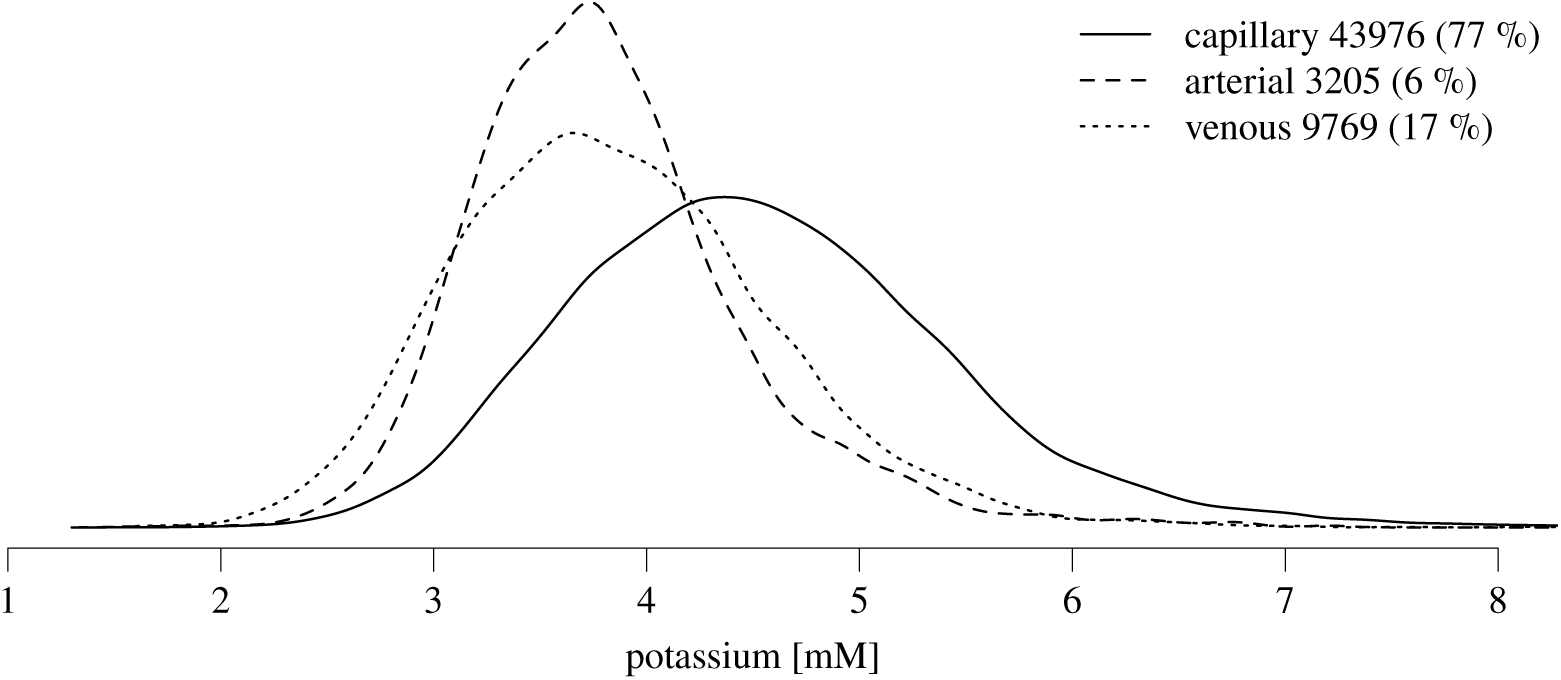
Plotted kernel density estimation of potassium values from reference population by sample type (arterial, venous, capillary). Percentages represent the share of the sum of all values included in this figure. While the shape is similiar, the location of the capillary sample distribution is considerably higher.

**Table 4:** Descriptive statistics of the distributions of potassium values from each source. The column with *M* [*P*_2.5_, *P*_97.5_] are the reference values brought forward in this manuscript. *n*, sample size, *x̂*, mode; *CrI*, 95 % mass credible interval; *x̅*, mean; *SD*, standard deviation; *γ*, Fisher-Pearson standardized moment coefficient of skewness.

| source | $n$ | $M[P_{2.5}, P_{97.5}]$ | $\hat{x}[CrI]$ | $\bar{x} \pm SD$ | $\gamma$ |
| --- | --- | --- | --- | --- | --- |
| *all chronological ages* |  |  |  |  |  |
| arterial | 3205 | 3.7[2.8, 5.3] | 3.7[2.8, 5.2] | $3.8 \pm 0.7$ | 1.31 |
| venous | 9769 | 3.8[2.6, 5.5] | 3.7[2.5, 5.3] | $3.9 \pm 0.8$ | 0.76 |
| capillary | 43976 | 4.5[3.0, 6.5] | 4.4[2.9, 6.3] | $4.5 \pm 0.9$ | 0.73 |
| *greater than 7<sup>th</sup> day of life* |  |  |  |  |  |
| arterial | 387 | 4.0[2.6, 5.9] | 4[2.6, 5.6] | $4.1 \pm 0.8$ | 1.28 |
| venous | 2995 | 4.0[2.6, 5.5] | 4.1[2.7, 5.6] | $4.0 \pm 0.8$ | 0.45 |
| capillary | 21949 | 4.5[3.0, 6.3] | 4.4[2.9, 6.1] | $4.5 \pm 0.9$ | 0.46 |

### Comparison of Capillary Potassium with Time-Matched Arterial or Venous Samples

Figure 3 depicts a two-dimensional kernel density estimator showing value pairs of capillary potassium measurements in relation to their difference from the nearest control, which come from arterial or venous samples drawn within a time interval of Δ*t* = *t*_capillary_ ± 2 h. The lines are exponentially cast isodenses labeled with their corresponding density value and represent the probability density of value pairs within that region.

**Figure 3:**
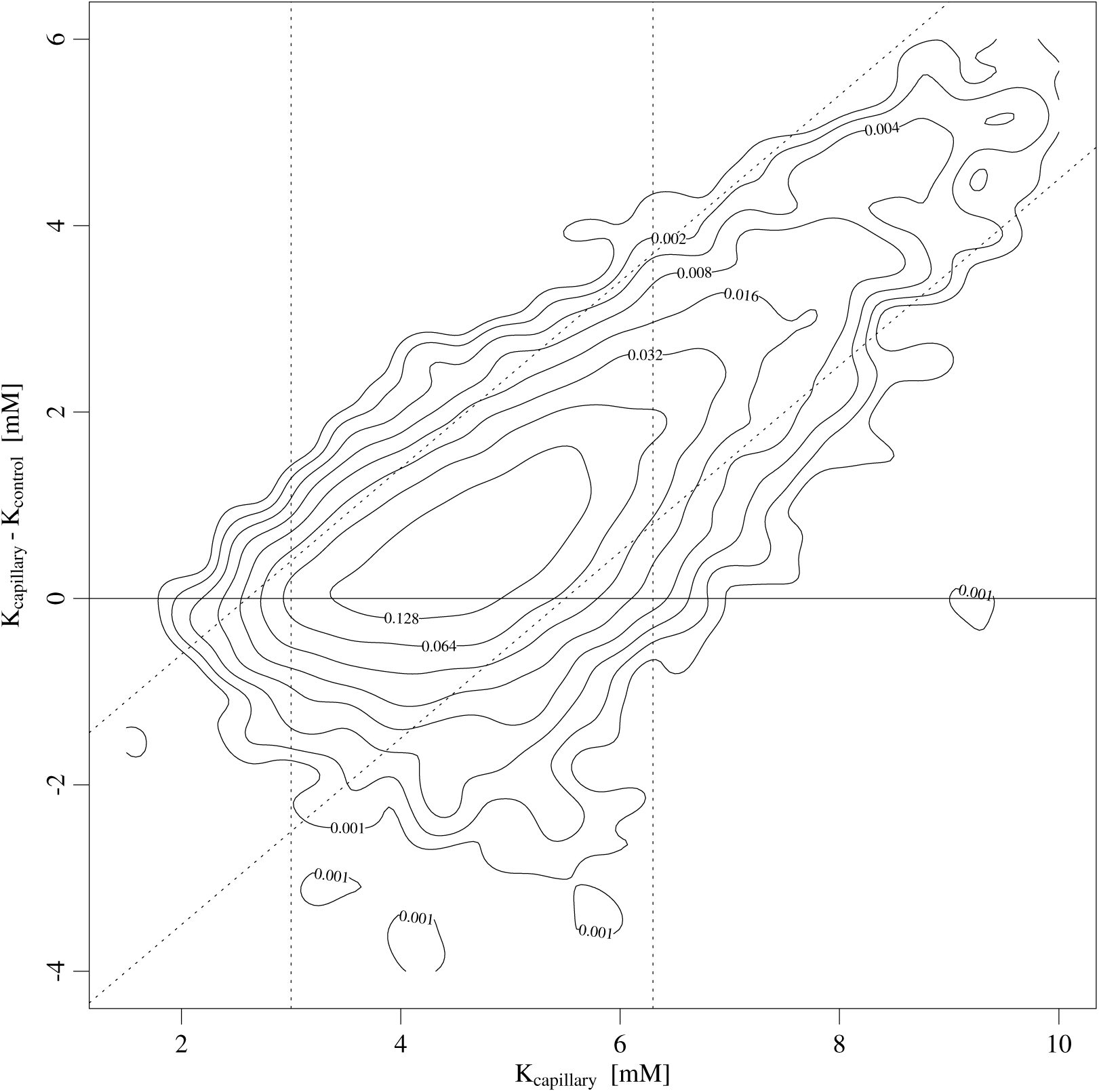
Contour plot of a two-dimensional kernel density estimation of the capillary potassium compared with its difference to the nearest time-matched control sample from another source (i. e. arterial or venous) within two hours *K*_control_. The exponentially cast isodensitiy lines are labelled with their corresponding density. Bandwidths were set and scaled automatically by kde2d using Silverman’s rule of thumb. The dashed lines represent the reference values after the 7^th^ day of life for venous and capillary samples from table 4. A total of N = 5403 such pairs are included in this plot.

The overall pattern shows a predominant cluster of data points. This suggests that, for most samples, capillary potassium levels are aligned with those from arterial or venous sources. The absolute differences are *K*_capillary_ − *K*_control_ = 0.7[−1.1, 3.6] meaning capillary values are higher. With increasing capillary values, the difference grows, with them becoming false-high almost exclusively compared to the controls above the upper limit of the reference interval of 6.3 from table 4.

Using this time-matched control sample the capillary interval *K*_cap_ = [3, 6.3] has 88 % sensitivity, 94 % specificity, 94 % positive predictive values, and 88 % negative predictive values when judging the controls to *K*_ven_ = [2.6, 5.5].

## Discussion

We report neonatal RI for potassium from integrated electronic patient records. To the best of our research we are the first to do this. Multivariate analysis gave reason to report separate RI for capillary samples due to their distinction. These capillary RI are notably higher. To test their clinical use, we compared capillary values with intra-individually time-matched arterial or venous samples using our new RI; We found that the RI are helpful in guiding clinical attention away from false-high capillary measurements. Usually, RI are derived from a population of healthy subjects. To avoid ethical dilemmata it has been suggested that values for children could be established indirectly, i. e. on existing large data sets by means of computation. It follows the notion, that the distribution of the healthy population can be derived from these data sets. [10] It is mostly done purely on the laboratory values and rarely external information has been put in. If so, it was information pertaining to the whole case like e. g. diagnoses without temporal resolution. [11]

Prematurity is a medical condition leading to various health issues arising at different times during the often long hospital stays. To account for them, we made use of a strength of our data set: Instead of excluding a severly ill patient totally, we excluded all samples that were taken during a severely impaired condition. To do this, we utilized the wide variety of connected data in our EPR. This may, at least in part, explain the differences to previously published RI for neonates:

While recruiting healthy participants directly from the community, the CALI-PER study used a different approach in infants. Samples before the 15^th^ day of life came from healthy neonates leaving maternity ward. From older infants leftover blood from outpatient clinics was used. [12] The resulting data were analyzed as outlined by the US-American Clinical and Laboratory Standards Institute. For each partition by sex and age 120 patients were sampled. Reference intervals were calculated from 131 infants to [3.9, 6.0] using a large automated analyzer [13] and for 275 subjects from birth to 18 years to [3.5, 4.7] using a blood gas analyzer. Sampling modality was not specified.

The PEDREF study reported indirect pediatric RI on 15 laboratory parame-ters by means of mining results from 13 laboratories which used different large automated analyzers [14]. If measurements were repeated within a certain time interval (for infants younger than 100 days within 50 day), values were filtered out. They arrived at 164 405 potassium values from ion-specific electrode mea-surements over all age groups after filtering out 82 % of the original sample. The publication contains no statements on age distribution or prematurity. It though seems reasonable to assume, that our sample size rivals that of PEDREF, espe-cially since the filtration criteria of PEDREF likely excluded all small preterm infants. The data for each day of life fed an optimization algorithm. It optimizes a Kolmogorov-Smirnov distance between a hypothetical Gaussian distribution and progressively truncated Box-Cox-transformed input data. [15] From the resulting data sets sex-specific LMS models were derived which can predict daily reference ranges. Between days of life 8 through 90 their LMS models predict daily reference ranges whose medians were 4.9[4.1, 5.8] for boys and 0.1 higher for girls. No destinction was made regarding sample type, disease, or short-term survival. The algorithm was further developed to eliminate the need for filtering or the specifica-tion of additional parameters. This enhanced version is publicly distributed as the `refineR` package for the R software environment. [16] When applied to our set of unfiltered capillary values without supplying any meta information, `refineR` returns an interval of [3.01, 6.25]. This is very close to our own results which are strongly based on the rich metadata in our database.

These pediatric RI studies pertain to chronological age. Prematurity, however, adds gestational age as a second dimension. In our multivariate analysis we found neither of them to be significant explanatory variables – our surrogate SIC and the date, on the other hand, were significant. New statistical methods allow to account for multivariate influences but to the best of our research they have not found implementation in laboratory medicine.

A narrative review by Chevalier is most commonly cited as source for potas-sium RI in the preterm of [4.5, 6.5]. [17] This was gleaned from three publications involving 64 preterm infants in total by delineating those who suffered complica-tions due to hyperkalemia such as cardiac arrest. In some of the cited sources the blood was stated as to come from non-capillary sources, others did not describe the sampling at all.

RI were published for cord blood of 591 neonates from all gestational age ranges currently considered viable. The RI were considerably broader for younger gestational ages being [2.7, 7.9] for preterm and [3.8, 5.7] for term infants. Gesta-tional age by cesarean delivery was significantly associated with lower potassium; gestational age alone, however, was not. [18]

Capillary sampling is a mainstay of clinical chemistry in neonates, as it helps avoid complications associated with vascular puncture, such as thrombosis and difficulty in cannulation. However, the validity of clinical chemistry derived from it has been called into question, this is especially true for potassium: Nowak identified 247 pairs of capillary and venous potassium samples drawn within ±4 h in older children admitted for acute gastroenteritis. [19] Agreement was deemed low with a mean absolute error of 0.51 ± 0.69 mM, our value was 0.83 ± 1.15 respectively. Applying the same RI for both sample types sensitivity calculated to 50 % and specificity to 88 %. When using a reference interval specific to the capillary sampling *K*_cap_ = [3, 6.3] we attain much higher test performance metrics. This interval performs acceptable in ruling out extreme actual potassium values.

Yang compared blood gas analyzes from 33 paired capillary and arterial sam-ples of preterm infants admitted to NICU. [20] Apart from the physiologically incomparable oxygen measurements, correlation was lowest for potassium.

Based on our findings, we would suggest further studies to include data from and validate with other centers.

Our study is retrospective and is thus prone to changes in practice. We assume these changes to reflect in the variable calendar date. Its influence in the multivari-ate analysis was stronger than all other variables apart from capillary sampling. Adding data of other centers would improve transferability of our RI. The rich variety of data in our sample has the potential to specifically inform other similar analyses for example of other laboratory parameters.

## Conclusion

The premature infant receives numerous painful interventions on a daily basis. [21] Their immature descending control of nociception makes pain feel more intensely. [22, 23] Further research into the topic of RI for the premature infant is thus direly needed to help clinicians with interpreting laboratory values and maybe thus forgoing of controls.

Hyperkalemia can bear important risks, but immediate resampling should only be done if clinical assessment of potassium intake, monitor ECG, or urine output are abnormal. [17] We hope that our new interval *K*_cap_ = [3, 6.3] will help in further reducing painful controls of the often spuriously increased capillary potassium.

## Data Availability

All data produced in the present study are available upon reasonable request to the authors.

## List of Acronyms

AIC: Akaike’s information criterion.
BGA: blood gas analysis.
BPD: bronchopulmonary dysplasia.
CLSI: Clinical & Laboratory Standards In-stitute.
CrI: credible interval.
ECG: electrocardiogram.
EPR: electronic patient record.
FiO_2_: fraction of inspired oxygen.
h.b.s.: hours before sampling.
IVH: intraventricular hemorrhage.
LMS: location, scale and shape.
M: median.
NEC: necrotizing enterocolitis.
NICU: neonatal intensive care unit.
pH: pondus hygrogenii.
RI: reference interval.
ROP: retinopathy of prematurity.
SD: standard deviation.
SDS: standard deviation score.
SIC: severly impaired condition.
SIP: spontaneous intestinal perforation.

## Acknowledgements

For preparing patient data are grateful for the services of theData IntegrationCenter of our hospital, which is funded by the German FederalMinistry of Education and Research (Grant No. 01KX2121).

## References

1. Defining, Establishing and Verifying Reference Intervals in the Clinical Laboratory 3rd ed. (ed CLSI) EP28-A3c. 60 pp. (Clinical and Laboratory Standards Institute, Wayne, PA).

2. Bergmann, K. E. et al. Ethische und rechtliche Aspekte der epidemiologischen Forschung mit Kindern und Jugendlichen in Deutschland am Beispiel des Kinder-und Jugendgesund-heitssurveys. Ethik in der Medizin 16, 22–36. doi:10.1007/s00481-004-0279-0.

3. Thierfelder, W. et al. Biochemische Messparameter im Kinder-und Jugendgesundheitssurvey (KiGGS). Bundesgesundheitsblatt 50, 757–770. doi:10.1007/s00103-007-0238-2.

4. Quante, M. et al. The LIFE Child Study: A Life Course Approach to Disease and Health. BMC Public Health 12, 1021. doi:10.1186/1471-2458-12-1021.

5. Allegaert, K. et al. The Publication Quality of Laboratory Values in Clinical Studies in Neonates. Pediatric Research. doi:10.1038/s41390-022-02385-1.

6. Radiometer. ABL800 FLEX Reference Manual (Brønshøj).

7. Sheather, S. J. & Jones, M. C. A Reliable Data-Based Bandwidth Selection Method for Kernel Density Estimation. Journal of the Royal Statistical Society. Series B (Methodological) 53, 683–690. https://www.jstor.org/stable/2345597.

8. Silverman, B. W. Density Estimation for Statistics and Data Analysis Monographs on Statistics and Applied Probability 26. 175 pp. https://ned.ipac.caltech.edu/level5/March02/Silverman/Silver_contents.html (Chapman & Hall/CRC, Boca Raton).

9. Meredith, M. & Kruschke, J. K. HDInterval: Highest (Posterior) Density Intervals ver-sion 0.2.4. https://cran.r-project.org/web/packages/HDInterval/index.html (2024).

10. Jones, G. R. et al. Indirect Methods for Reference Interval Determination – Review and Recommendations. Clinical Chemistry and Laboratory Medicine 57, 20–29. doi:10.1515/cclm-2018-0073.

11. Poole, S., Schroeder, L. F. & Shah, N. An Unsupervised Learning Method to Identify Reference Intervals from a Clinical Database. Journal of Biomedical Informatics 59, 276–284. doi:10.1016/j.jbi.2015.12.010.

12. Colantonio, D. A. et al. Closing the Gaps in Pediatric Laboratory Reference Intervals: A CALIPER Database of 40 Biochemical Markers in a Healthy and Multiethnic Population of Children. Clinical Chemistry 58, 854–868. doi:10.1373/clinchem.2011.177741.

13. Tahmasebi, H. et al. Pediatric Reference Intervals for Clinical Chemistry Assays on Siemens ADVIA XPT/1800 and Dimension EXL in the CALIPER Cohort of Healthy Children and Adolescents. Clinica Chimica Acta 490, 88–97. doi:10.1016/j.cca.2018.12.011.

14. Zierk, J. et al. High-Resolution Pediatric Reference Intervals for 15 Biochemical Analytes Described Using Fractional Polynomials. Clinical Chemistry and Laboratory Medicine (CCLM) 59, 1267–1278. doi:10.1515/cclm-2020-1371.

15. Zierk, J. et al. Reference Interval Estimation from Mixed Distributions Using Truncation Points and the Kolmogorov-Smirnov Distance (Kosmic). Scientific Reports 10, 1704. doi:10.1038/s41598-020-58749-2.

16. Ammer, T. et al. refineR: A Novel Algorithm for Reference Interval Estimation from Real-World Data. Scientific Reports 11, 16023. doi:10.1038/s41598-021-95301-2.

17. Chevalier, R. L. What Are Normal Potassium Concentrations in the Neonate? What Is a Reasonable Approach to Hyperkalemia in the Newborn with Normal Renal Function? Seminars in Nephrology 18, 360–361.

18. Stritzke, A. et al. Cord-Blood Derived Chemistry Reference Values in Preterm Infants for Sodium, Chloride, Potassium, Glucose, and Creatinine. American Journal of Perinatology 41, 722–729. doi:10.1055/a-1730-8536.

19. Nowak, J. K. et al. Reliability of Capillary Blood Potassium Measurements in Children with Acute Gastroenteritis. Archives of Disease in Childhood 103, 1091–1093. doi:10.1136/archdischild-2017-314561.

20. Yang, K.-C. et al. The Comparison between Capillary Blood Sampling and Arterial Blood Sampling in an NICU. Acta Paediatrica Taiwanica = Taiwan Er Ke Yi Xue Hui Za Zhi 43, 124–126.

21. Simons, S. H. P. et al. Do We Still Hurt Newborn Babies? Archives of Pediatrics & Adolescent Medicine 157, 1058. doi:10.1001/archpedi.157.11.1058.

22. Walker, S. M., Beggs, S. & Baccei, M. L. Persistent Changes in Peripheral and Spinal Nociceptive Processing after Early Tissue Injury. Experimental Neurology 275, 253–260. doi:10.1016/j.expneurol.2015.06.020.

23. Hatfield, L. A. Neonatal Pain: What’s Age Got to Do with It? Surgical Neurology International 5, 479. doi:10.4103/2152-7806.144630.

